# SARS-CoV-2 seroprevalence and gender-related haematological features in asymptomatic blood donors

**DOI:** 10.1101/2021.04.28.21256242

**Authors:** M Carmen Martín, M Isabel González, Nuria Holgado, Ana I Jimenez, Nuria Ortega, Isabel Page, Alba Parrado, María Pérez, Lydia Blanco-Peris

**Author notes:** **Corresponding author**: M Carmen Martín; Centro de Hemoterapia y Hemodonación de Castilla y León, Paseo de Filipinos s/n, 47007 Valladolid, Spain.

## Abstract

**Background and objectives:** COVID-19 can either cause death or go unnoticed but antibodies will remain protecting us of SARS-CoV-2 reinfection for an uncertain time and to an uncertain extent. Our aim was to describe seroprevalence evolution from summer 2019 to autumn 2020 in Spain and to describe its relationship with age, blood group and haematological parameters.

**Materials and methods:** Sera and plasma from historical donation archives excluding convalescent were randomized and a total of 12,313 donations tested by a Chemiluminiscent analysis for anti SARS-CoV-2 N protein total immunoglobulins. Blood donors were 60.9% males, average age 46+/-13. Sex, age, blood group, blood cell counts and percentages and immunoglobulin concentrations were extracted from electronic recordings.

**Results:** A seroprevalence of 6.7% in blood donors was found by the end of the first wave. No differences by sex, age or blood group were found regarding antibodies. Leukocyte count (p=0.026), haematocrit (p<0.001) and haemoglobin (p<0.001) were lower in positive donations than in negative ones. Sex differences were present in neutrophils, leukocytes, haemoglobin and haematocrit as related to SARS-CoV-2 antibodies.

**Conclusions:** Seroprevalence due to asymptomatic cases would resemble that of global population. Sex and age would not affect COVID-19 susceptibility but its severity. Gender differences related to COVID-19 in leukocytes, haemoglobin and haematocrit would be present in asymptomatic individuals. Further studies are needed to confirm these gender differences as they can help better understand the immune response to COVID-19, its pathogenesis and prognosis.

## Introduction

An unknown amount of individuals infected with SARS-CoV-2 present no symptoms or a mild disease that can go unnoticed. This make it difficult to estimate infection rates, prevalence, calculating absolute risks of COVID-19 or predicting the spread of the virus and the growth of herd immunity. [1]. Immunization either due to infection or to vaccination leads specific T cell responses and production of antibodies against SARS-CoV-2 usually peaking 8–15 days after infection [2]. Antibodies that neutralize the virus are the primary protection against COVID-19.

Antibody assays are quite different: they either detect antibodies against different viral proteins (S1, S1/S2, RBD or NC) or different immunoglobulin classes: IgG, IgM, IgA or their combinations. It should be noticed that every approved vaccine in Europe to date (May 2020) enhances anti-S responses, whereas an infection leads to a multi specific response to SARS-CoV2 antigens. To make it even messier, many factors can influence test performance, including cross-reactivity with other coronaviruses or platform (laboratory-based vs point-of-care, lateral flow). Chemiluminiscence assays have suitable performances regarding both sensibility and sensitivity while correlation to neutralizing antibodies would be around 0.7 [3].

Our region, Castilla y Leon, with a population of 2.299.598 inhabitants, has accumulated nearly 225.000 COVID-19 confirmed cases (https://analisis.datosabiertos.jcyl.es/), with an overall case-fatality rate of 3.01 to date. Our institution, Centro de Hemoterapia de Castilla y León keeps within its Biobank plasma samples from every single donation along the last ten years, that makes up around one million samples. Any demographic data such as sex or age and laboratory parameters (blood counts, blood group, haemoglobin, etc.…) is as well kept in a database (Hemasoft, eDelphyn).

Blood donor-based serosurveillance is a powerful and cost-effective strategy to monitor infectious diseases. There are quite a lot of infections for which routine donor screening is undoubtedly useful, including human immunodeficiency virus (HIV), hepatitis C virus (HCV), hepatitis B virus (HBV), human T-lymphotrophic virus (HTLV), and West Nile virus (WNV) [1], babesiosis or Crimea-Congo fever. The use of blood donor samples means we are sampling mainly asymptomatic and recovered cases of COVID-19 (donation is allowed after 28 days following COVID-19 resolution of symptoms).

Seroprevalence surveys are then needed to monitor coronavirus pandemic, both before and after vaccination strategies [4]. Reported COVID-19 cases do not represent the full SARS-CoV-2 disease burden due to low detection rates, especially at the beginning of pandemics. Case reports are dependent on patients’ seeking health care, massive local screenings or regional tracking activities. Analysis of data from seroprevalence and serosurveillance is a common strategy for estimating underreporting and real disease burden. Time between infection and antibody development or antibody waning must be considered to understand seroprevalence surveys avoiding biases. Serological surveys with a random sampling design of the general population are difficult to perform in a pandemic, but they would yield a seroprevalence estimate least likely to be affected by selection bias. Blood donors are a quite representative subset of general population aged 18-65 and our historical plasma collection was big enough to perform a systematic time-layered randomization.

Heterogeneity of susceptibility and transmission are hard to evaluate [5]. A portion of the population is not susceptible to infection from the first pathogen contact. Some of them may have pre-existing immunity via cross-reactivity or particular host factors such as mucosal immunity or trained innate immunity protection (as it has been reported to be conferred by DTP or BCG vaccination)[6]. There is as well a proportion of seronegative individuals that will develop immunity by T cell mediated responses without exhibiting an antibody response [7].

Our starting hypothesis was the existence of a certain number of asymptomatic carriers of the SARS-CoV-2 virus that would develop antibodies against it [7] and subsequently contribute to herd immunity. Those asymptomatic cases were unnoticeable before SARS-CoV-2 testing began and might had been circulating from an unknown moment. Early cases have been described in France [8], arising the question of whether the pathogen could have been circulating before the official recording of the first cases.

The main aim of this study was to determine which percentage of the population has had contact with SARS-CoV-2 at different times and had therefore developed antibodies against it. A secondary goal was to describe related haematological features of seropositive donors and establish whether sex, age, blood group or haematological abnormalities could be related to past COVID-19 infection.

Knowledge of COVID-19 epidemiological and haematological features should help make a good forecast of resources for possible future outbreaks or vaccination strategies, and would facilitate decisions in the social sphere by a more reliable estimate of the percentage of immunized individuals.

## MATERIAL and METHODS

Seroprevalence and haematological features were studied blood, plasma (excluding convalescent) and platelet donations from July 2019 to October 2020. The 101,183 donations from 70,181 donors collected since 19/07/2019 (week −24) to 19/10/2020 (week 43) were systematically randomized to select a minimum of 127 donations per week, calculated on the basis of the total number of donations and the length of the analysis period. Then, 1650 convalescent plasma donation were excluded. A total of 12,718 samples of 12,313 donations from 11,444 donors over 18 years old were included. All haematological and demographic data were extracted from our electronic database eDelphyn (Hemasoft). Variables analysed included age, sex, blood group, and laboratory data: leukocyte (WBC), neutrophil, lymphocyte, platelet, monocyte, eosinophil and basophil counts (cells* 10^3^/µL) and their percentages, serum immunoglobulins IgG, IgA and IgM (mg/dL), haemoglobin (Hb), and haematocrit (HCT) were analysed as well.

An automated chemiluminiscence double-antigen sandwich immunoassay for the in vitro semi quantitative detection of total antibodies to SARS-CoV-2 in human plasma and serum frozen samples. The target antigen of this immunoassay is a recombinant nucleocapsid (N) protein. Elecsys® Anti-SARS-CoV-2 (Roche, Basel, Switzerland) detects antibodies correlating with virus-neutralizing ones and is therefore useful to help characterize the immune reaction to SARS-CoV-2 [9, 10]. Immunoassay was validated by our serology lab by testing of 6 pairs of samples (plasma EDTA and serum) from diagnosed PCR-positive, symptomatic cases infected by mid-April, that were previously reported positive by the Spanish National Microbiology Centre, and checked to be as well positive for IgG (Chemiluminiscence, N protein, Abbott Alinity S, Chicago USA) and IgA (ELISA, S protein, Euroimmunn, Lübeck, Germany) antiSARS-CoV-2. Another set of ten prepandemic samples, therefore supposed to be negative, were equally analysed. 405 donations were analysed both in serum and plasma to verify interchangeability. A 100% concordance was yielded by all these validation assays. The cut-off was that recommended by manufacturer (OD>1 to report reactivity). Researchers performing anti SARS-CoV-2 analyses were blind to the condition of COVID-19 convalescence and to any other characteristic of the donors or to the donation dates.

Demographic and clinical characteristics of patients are expressed as their mean, median, standard deviation (SD) and interquartile range (IQR) for continuous variables and frequency distributions are reported for categorical variables. Age was analysed both as continuous and categorical variable; in the latter case was recoded into 4 groups: <30, 30-45, 45-60 and 60-75 years old.

Kolmogorov-Smirnov test was performed on each continuous variable to contrast normality. Only IgG and IgM serum levels followed normal distributions, therefore non-parametric Mann-Whitney U test was performed. To contrast independence of categorical variables, Pearson’s Chi-square and Fisher’s exact test were carried out. All tests were calculated with a confidence level of 0.05.

The Biobank is included in the National Registry of Biobanks (RD17 / 16/2011) with the number B.0000264. The institution holds an ISO 9001: 2015 certification endorsing our granting of safety and traceability of any human biological sample we distribute, always behaving Spanish and European rules on human samples and data protection management.

This study was conducted according with national regulations, institutional policies and in the tenets of the Helsinki Declaration. It was approved by the Valladolid Health Area Drug Research Ethics Committee, on June 11th, 2020 with the reference number BIO-2020-93. Included donors consented to participate in Biobank research activities. The privacy rights were always observed.

## Results

The 101,183 donations from 70,181 donors along 67 weeks (July 2019 to October 2020) were systematically randomized, discarding convalescent plasma ones and a total of 12,313 donations (either whole blood, plasmapheresis or platelet apheresis) were tested for total anti SARS-CoV-2 antibodies. Donors were 11,444 individuals, 60.91% males, aged 18-75, average 46+/-13, 45.8% A, 40.3% O, 10.56% B, 3.31% AB. Baseline characteristics descriptive analyses are summarized in Table 1. Immunoglobulin results are usually performed only once a year in plasma donors.

**Table 1.** Baseline characteristics of donations.

|  | n | mean | median | SD | IQR |
| --- | --- | --- | --- | --- | --- |
| Age | 12313 | **44,0** | **45,8** | 12,8 | 34,2-54,3 |
| IgG | 199 | **1066,9** | **1038,0** | 217,8 | 910-1185 |
| IgA | 199 | **236,6** | **224,0** | 100,9 | 162-292 |
| IgM | 199 | **113,0** | **100,0** | 64,3 | 69-138 |
| Leukocyte | 10105 | **7,4** | **7,2** | 1,8 | 6,1-8,5 |
| Haemoglobin | 11604 | **14,9** | **14,9** | 1,3 | 14-15,8 |
| Haematocrit | 11604 | **45,1** | **45,1** | 3,5 | 42,6-47,5 |
| Platelets | 11603 | **247,9** | **243,0** | 54,3 | 211-280 |
| Neutrophil count | 11603 | **4,4** | **4,2** | 1,4 | 3,4-5,2 |
| Neutrophil % | 11603 | **58,8** | **58,9** | 7,5 | 53,8-63,9 |
| Lymphocyte count | 11602 | **2,3** | **2,3** | 0,7 | 1,9-2,7 |
| Lymphocyte % | 11602 | **31,7** | **31,5** | 6,9 | 27-36,2 |
| Monocyte count | 11602 | **0,5** | **0,5** | 0,1 | 0,4-0,6 |
| Monocyte% | 11602 | **6,5** | **6,4** | 1,5 | 5,5-7,4 |
| Eosinophil count | 11603 | **0,2** | **0,2** | 0,1 | 0,1-0,2 |
| Eosinophil % | 11603 | **2,5** | **2,1** | 1,8 | 1,3-3,2 |
| Basophil count | 11603 | **0,0** | **0,0** | 0,0 | 0-0 |
| Basophil % | 11603 | **0,5** | **0,5** | 0,2 | 0,3-0,6 |

No differences in anti SARS-CoV2 reactivity due to sex, age or blood group of donations were found (Table 2).

**Table 2.** Sex, age range and blood group of positive/negative anti SARS-COV2 donations analysed.

|  | Negative |  | Positive |  |
| --- | --- | --- | --- | --- |
|  | n | % | n | % |
| Female | 4531 | 38.1% | 164 | 40.3% |
| Male | 7375 | 61.9% | 243 | 59.7% |
| <30 | 3551 | 29.8% | 104 | 25.6% |
| 30 to 45 | 5036 | 42.3% | 180 | 44.2% |
| 45 to 60 | 1151 | 9.7% | 42 | 10.3% |
| 60 to 75 | 2168 | 18.2% | 81 | 19.9% |
| A | 5411 | 45.5% | 198 | 48.6% |
| AB | 423 | 3.6% | 16 | 3.9% |
| B | 919 | 7.7% | 22 | 5.4% |
| O | 5151 | 43.3% | 171 | 42.0% |

Three 2019 donations were reactive (from July, September and October; 1,142 donations from year 2019 were analyzed) but none of them were positive when testing separately IgG, IgA or IgM. Their ODs were 1.05, 1.08 and 5.46 respectively.

Seropositivity rate (positive donations x 100 / total analyzed donations per week) grew up from week 11 to week 25, reaching a 11.1% peak, with just subtle descents until week 43 when the second wave arose in our country. Plateau was reached by the last week of May 2020, 11 weeks after close lockdown in Spain and 17 weeks after 31/01, the date of the first declared case in or country. 660 out of 9,886 single donors were positive within weeks 11 to 43 (Fig 1). Seroprevalence was therefore 6.73% at the end of the first wave.

**Fig 1.**
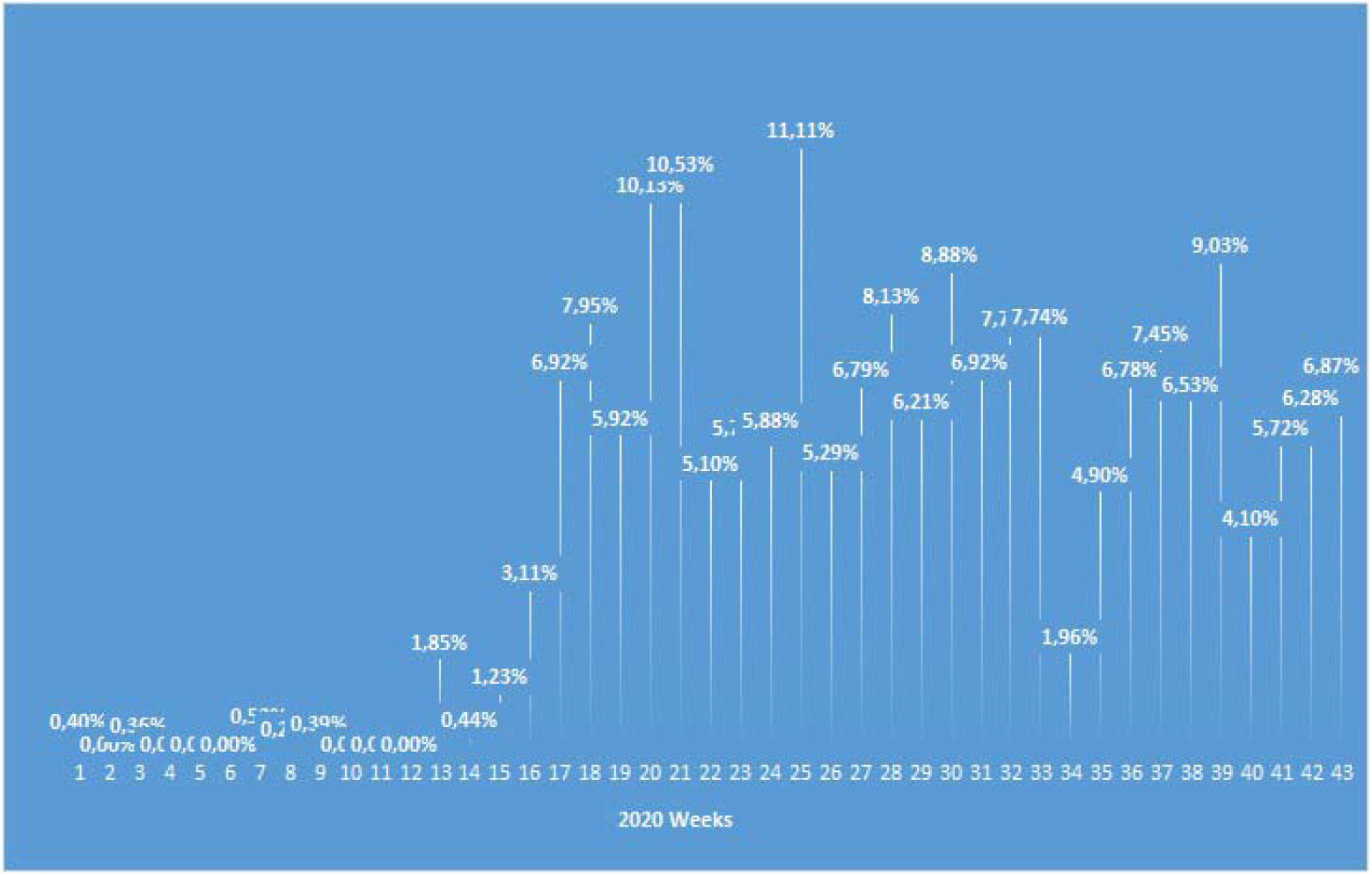
COVID-19 seropositive donation rate. 2020 evolution by weeks.

Immunoglobulin levels didn’t show any differences as comparing positive and negative donations (Table 3). Haemoglobin (14.9 vs 14.7 g/dL; p<0.001) and haematocrit (45.2% vs 44.4%; p<0.001) were both significantly lower in positive donations. Leukocyte count (WBC) was as well slightly lower (7.23 vs 7.06 cells* 10^3^/µL; p=0.026). Trends were found of lower number of neutrophils (4.3 vs. 4.2 cells* 10^3^/µL; p=0.117), lower count of monocytes (0.46 vs. 0.45 cells* 10^3^/µL; p=0.117) and higher percent of basophils (0.49% vs 0.51%; p=0.131) in positive donations as compared with negative ones.

**Table 3.** Descriptive analyses of age, immunoglobulins and haemogram of reactive and non-reactive donations.

reactive and non-reactive donations
|  |  | n | mean | median | SD | IQR |
| --- | --- | --- | --- | --- | --- | --- |
| Age | Negative | 11906 | 44.0 | **45.8** | 12.8 | 34.2-54.2 |
|  | Positive | 407 | 44.2 | **46.5** | 13.0 | 32.8-54.8 |
| IgA | Negative | 194 | 236.9 | **223.5** | 101.3 | 163-292 |
|  | Positive | 5 | 224.4 | **274.0** | 92.3 | 134-282 |
| IgG | Negative | 194 | 1065.4 | **1037.5** | 217.4 | 910-1185 |
|  | Positive | 5 | 1123.8 | **1095.0** | 249.9 | 924-1229 |
| IgM | Negative | 194 | 113.3 | **100.5** | 64.8 | 69-138 |
|  | Positive | 5 | 103.0 | **89.0** | 41.7 | 78-97 |
| WBC<sup>a</sup> | Negative | 9759 | 7.4 | **7.2** | 1.8 | 6.13-8.49 |
|  | Positive | 346 | 7.2 | **7.1** | 1.6 | 6.11-8.12 |
| Hb <sup>b</sup> | Negative | 11218 | 14.9 | **14.9** | 1.3 | 14-15.8 |
|  | Positive | 386 | 14.7 | **14.7** | 1.2 | 13.8-15.4 |
| Hematocrite <sup>b</sup> | Negative | 11218 | 45.1 | **45.2** | 3.5 | 42.7-47.6 |
|  | Positive | 386 | 44.5 | **44.4** | 3.5 | 42.3-46.6 |
| Platelets | Negative | 11217 | 247.9 | **243.0** | 54.3 | 211-280 |
|  | Positive | 386 | 247.3 | **242.5** | 55.7 | 206-276 |
| Neutrophil |  |  |  |  |  |  |
| count <sup>c</sup> | Negative | 11217 | 4.4 | **4.3** | 1.4 | 3.44-5.19 |
|  | Positive | 386 | 4.3 | **4.2** | 1.2 | 3.34-5.03 |
| Neutrophil % | Negative | 11217 | 58.8 | **58.9** | 7.5 | 53.8-63.9 |
|  | Positive | 386 | 58.5 | **58.7** | 7.1 | 54.3-63.4 |
| Lymphocyte |  |  |  |  |  |  |
| count | Negative | 11216 | 2.3 | **2.3** | 0.7 | 1.86-2.72 |
|  | Positive | 386 | 2.3 | **2.2** | 0.6 | 1.89-2.64 |
| Lymphocyte % | Negative | 11216 | 31.7 | **31.5** | 6.9 | 27-36.2 |
|  | Positive | 386 | 32.1 | **32.4** | 6.6 | 27.3-36.1 |
| Monocyte |  |  |  |  |  |  |
| count <sup>d</sup> | Negative | 11216 | 0.5 | **0.46** | 0.1 | 0.38-0.56 |
|  | Positive | 386 | 0.5 | **0.46** | 0.1 | 0.37-0.54 |
| Monocyte % | Negative | 11216 | 6.5 | **6.4** | 1.5 | 5.5-7.4 |
|  | Positive | 386 | 6.5 | **6.4** | 1.4 | 5.5-7.3 |
| Eosinophil |  |  |  |  |  |  |
| count | Negative | 11217 | 0.2 | **0.2** | 0.1 | 0.09-0.23 |
|  | Positive | 386 | 0.2 | **0.1** | 0.1 | 0.09-0.22 |
| Eosinophil % | Negative | 11217 | 2.5 | **2.1** | 1.8 | 1.3-3.2 |
|  | Positive | 386 | 2.4 | **2.0** | 1.7 | 1.3-3.1 |
| Basophil count | Negative | 11217 | 0.0 | **0.0** | 0.0 | 0.02-0.04 |
|  | Positive | 386 | 0.0 | **0.0** | 0.0 | 0.02-0.04 |
| Basophil % | Negative | 11217 | 0.5 | **0.5** | 0.2 | 0.3-0.6 |
|  | Positive | 386 | 0.5 | **0.5** | 0.3 | 0.4-0.6 |

Units: Immunoglobulins (mg/dL); Hb (g/dL); haemogram counts
(cells\*10<sup>3</sup>/μL)
<sup>a</sup> p=0.026
<sup>b</sup> p<0.001
<sup>c</sup> p=0.117
<sup>d</sup>p=0.131

As layering data by sex (Table 4), the fact arose that haematocrit (46.8% vs 45.9%; p<0.001) and haemoglobin (15.5 vs 15.3 g/dL; p<0.001) were significantly lower just in males and that positive females had significantly less WBC (7.67 vs 7.25 cells* 10^3^/µL; p=0.027) and neutrophil numbers (4.53 vs 4.27 cells* 10^3^/µL; p=0.014).

**Table 4.** Descriptive analysis of age, age, immunoglobulins and haemogram of reactive and non-reactive donations layered by sex.

|  |  |  | n | mean | median | SD | IQR |
| --- | --- | --- | --- | --- | --- | --- | --- |
| Age | Female | Negative | 4531 | 42.0 | **43.50** | 13.3 | 30.3-53.3 |
|  |  | Positive | 164 | 42.1 | **45.39** | 13.6 | 28.6-53.1 |
|  | Male | Negative | 7375 | 45.2 | **46.81** | 12.2 | 36.7-54.8 |
|  |  | Positive | 243 | 45.7 | **46.98** | 12.5 | 35.5-55.9 |
| IgA | Female | Negative | 54 | 214.3 | **200.00** | 97.0 | 157-250 |
|  |  | Positive | 3 | 235.3 | **274.00** | 105.5 | 116-316 |
|  | Male | Negative | 140 | 245.6 | **233.50** | 101.9 | 176.5-302.5 |
|  |  | Positive | 2 | 208.0 | **208.00** | 104.7 | 134-282 |
| IgG | Female | Negative | 54 | 1076.4 | **1062.50** | 191.3 | 964-1186 |
|  |  | Positive | 3 | 1082.7 | **1095.00** | 152.9 | 924-1229 |
|  | Male | Negative | 140 | 1061.2 | **1029.00** | 227.2 | 903-1181.5 |
|  |  | Positive | 2 | 1185.5 | **1185.50** | 436.3 | 877-1494 |
| IgM | Female | Negative | 54 | 143.1 | **129.50** | 64.6 | 94-169 |
|  |  | Positive | 3 | 114.3 | **89.00** | 53.7 | 78-176 |
|  | Male | Negative | 140 | 101.8 | **90.00** | 61.3 | 64.5-121 |
|  |  | Positive | 2 | 86.0 | **86.00** | 15.6 | 75-97 |
| Mean |  |  |  |  |  |  |  |
| corpuscular |  |  |  |  |  |  |  |
| volume | Female | Negative | 3618 | 91.2 | **91.50** | 4.8 | 88.4-94.4 |
|  |  | Positive | 142 | 91.2 | **91.40** | 4.1 | 88.9-93.5 |
|  | Male | Negative | 6141 | 90.9 | **91.00** | 4.6 | 88.1-93.8 |
|  |  | Positive | 204 | 90.6 | **91.00** | 4.8 | 88-94.1 |
| WBC | Female<sup>a</sup> | Negative | 3618 | 7.8 | **7.69** | 1.9 | 6.49-9 |
|  |  | Positive | 142 | 7.4 | **7.21** | 1.5 | 6.27-8.26 |
|  | Male | Negative | 6949 | 7.2 | **7.02** | 1.8 | 5.99-8.2 |
|  |  | Positive | 229 | 7.1 | **6.98** | 1.6 | 6.01-8.03 |
| Hb | Female | Negative | 4269 | 13.8 | **13.80** | 0.9 | 13.2-14.4 |
|  |  | Positive | 157 | 13.7 | **13.80** | 0.9 | 13.2-14.3 |
|  | Male<sup>b</sup> | Negative | 6949 | 15.5 | **15.50** | 1.0 | 14.9-16.2 |
|  |  | Positive | 229 | 15.3 | **15.30** | 1.0 | 14.7-15.8 |
| Haematocrit | Female | Negative | 4269 | 42.4 | **42.30** | 2.7 | 40.6-44 |
|  |  | Positive | 157 | 42.1 | **42.00** | 2.5 | 40.5-43.6 |
|  | Male<sup>b</sup> | Negative | 6949 | 46.8 | **46.80** | 2.8 | 45-48.6 |
|  |  | Positive | 229 | 46.2 | **45.90** | 3.0 | 44.3-47.7 |
| Platelets | Female<sup>c</sup> | Negative | 4268 | 265.9 | **261.00** | 56.6 | 228-300 |
|  |  | Positive | 157 | 261.0 | **255.00** | 60.5 | 219-296 |
|  | Male | Negative | 6949 | 236.8 | **233.00** | 49.6 | 204-266 |
|  |  | Positive | 229 | 237.9 | **234.00** | 50.2 | 203-266 |
| Neutrophil |  |  |  |  |  |  |  |
| count | Female | Negative | 4268 | 4.7 | **4.53** | 1.4 | 3.65-5.47 |
|  |  | Positive | 157 | 4.4 | **4.27** | 1.2 | 3.56-5.08 |
|  | Male | Negative | 6949 | 4.3 | **4.10** | 1.4 | 3.34-4.95 |
|  |  | Positive | 229 | 4.2 | **4.10** | 1.2 | 3.24-4.97 |
| Neutrophil |  |  |  |  |  |  |  |
| % | Female | Negative | 4268 | 59.2 | **59.40** | 7.4 | 54.2-64.3 |
|  |  | Positive | 157 | 58.1 | **58.10** | 7.2 | 54-62.9 |
|  | Male | Negative | 6949 | 58.6 | **58.70** | 7.5 | 53.6-63.6 |
|  |  | Positive | 229 | 58.8 | **58.70** | 7.0 | 54.6-63.5 |
| Lymphocyte |  |  |  |  |  |  |  |
| count | Female | Negative | 4267 | 2.5 | **2.39** | 0.7 | 1.97-2.88 |
|  |  | Positive | 157 | 2.4 | **2.35** | 0.6 | 2.02-2.81 |
|  | Male | Negative | 6949 | 2.2 | **2.18** | 0.6 | 1.8-2.62 |
|  |  | Positive | 229 | 2.2 | **2.17** | 0.6 | 1.8-2.51 |
| Lymphocyte |  |  |  |  |  |  |  |
| % | Female<sup>e</sup> | Negative | 4267 | 32.0 | **31.70** | 6.9 | 27.4-36.5 |
|  |  | Positive | 157 | 33.0 | **33.10** | 7.0 | 28.4-37 |
|  | Male | Negative | 6949 | 31.5 | **31.30** | 6.9 | 26.8-36.1 |
|  |  | Positive | 229 | 31.4 | **31.70** | 6.3 | 26.7-35.6 |
| Monocyte |  |  |  |  |  |  |  |
| count | Female<sup>f</sup> | Negative | 4267 | 0.5 | **0.46** | 0.1 | 0.37-0.55 |
|  |  | Positive | 157 | 0.5 | **0.44** | 0.1 | 0.36-0.52 |
|  | Male | Negative | 6949 | 0.5 | **0.46** | 0.1 | 0.38-0.56 |
|  |  | Positive | 229 | 0.5 | **0.47** | 0.1 | 0.38-0.55 |
| Monocyte |  |  |  |  |  |  |  |
| % | Female | Negative | 4267 | 6.1 | **6.00** | 1.4 | 5.1-6.9 |
|  |  | Positive | 157 | 6.1 | **6.00** | 1.4 | 5.2-6.9 |
|  | Male | Negative | 6949 | 6.7 | **6.60** | 1.5 | 5.7-7.6 |
|  |  | Positive | 229 | 6.7 | **6.70** | 1.4 | 5.8-7.6 |
| Eosinophil |  |  |  |  |  |  |  |
| count | Female | Negative | 4268 | 0.2 | **0.14** | 0.1 | 0.09-0.21 |
|  |  | Positive | 157 | 0.2 | **0.13** | 0.1 | 0.09-0.19 |
|  | Male | Negative | 6949 | 0.2 | **0.16** | 0.1 | 0.1-0.24 |
|  |  | Positive | 229 | 0.2 | **0.15** | 0.1 | 0.09-0.23 |
| Eosinophil |  |  |  |  |  |  |  |
| % | Female | Negative | 4268 | 2.2 | **1.80** | 1.6 | 1.2-2.8 |
|  |  | Positive | 157 | 2.3 | **1.80** | 1.7 | 1.1-2.8 |
|  | Male | Negative | 6949 | 2.7 | **2.20** | 1.8 | 1.4-3.4 |
|  |  | Positive | 229 | 2.5 | **2.10** | 1.7 | 1.4-3.2 |
| Basophil |  |  |  |  |  |  |  |
| count | Female | Negative | 4268 | 0.0 | **0.03** | 0.0 | 0.02-0.05 |
|  |  | Positive | 157 | 0.0 | **0.04** | 0.0 | 0.02-0.05 |
|  | Male | Negative | 6949 | 0.0 | **0.03** | 0.0 | 0.02-0.04 |
|  |  | Positive | 229 | 0.0 | **0.03** | 0.0 | 0.02-0.04 |
| Basophil % | Female<sup>g</sup> | Negative | 4268 | 0.5 | **0.40** | 0.2 | 0.3-0.6 |
|  |  | Positive | 157 | 0.5 | **0.50** | 0.3 | 0.3-0.6 |
|  | Male | Negative | 6949 | 0.5 | **0.50** | 0.2 | 0.3-0.6 |
|  |  | Positive | 229 | 0.5 | **0.50** | 0.2 | 0.4-0.6 |

\*p<0.05
Units:immunoglobulins (mg/dL); Hb(g/dL); haemogram counts
(cells\*10<sup>3</sup>/μL) and percent
<sup>a</sup>p=0.027
<sup>b</sup>p<0.001
<sup>c</sup>p=0.152
<sup>d</sup>p=0.065
<sup>e</sup>p=0.074
<sup>f</sup>p=0.132
<sup>g</sup>p=0.060

Some trends were found as well related to sex. Regarding positive women neutrophil percent would be lower (59.4% vs 58.10% p=0.065), lymphocyte percent, higher (31.7% vs 33.10%; p=0.074), platelet count, (261 vs 255 cells* 10^3^/µL; p=0.152), monocyte count (0.46 vs 0.44 cells* 10^3^/µL; p=0.132) and basophil percent (0.5% vs 0.4%; p=0.060) lower. Anaemia was more frequent in positive male donors as (6.1% vs 9.6%; p=0.031). No other significant differences were found either in males, females or overall (Fig 2).

**Fig 2.**
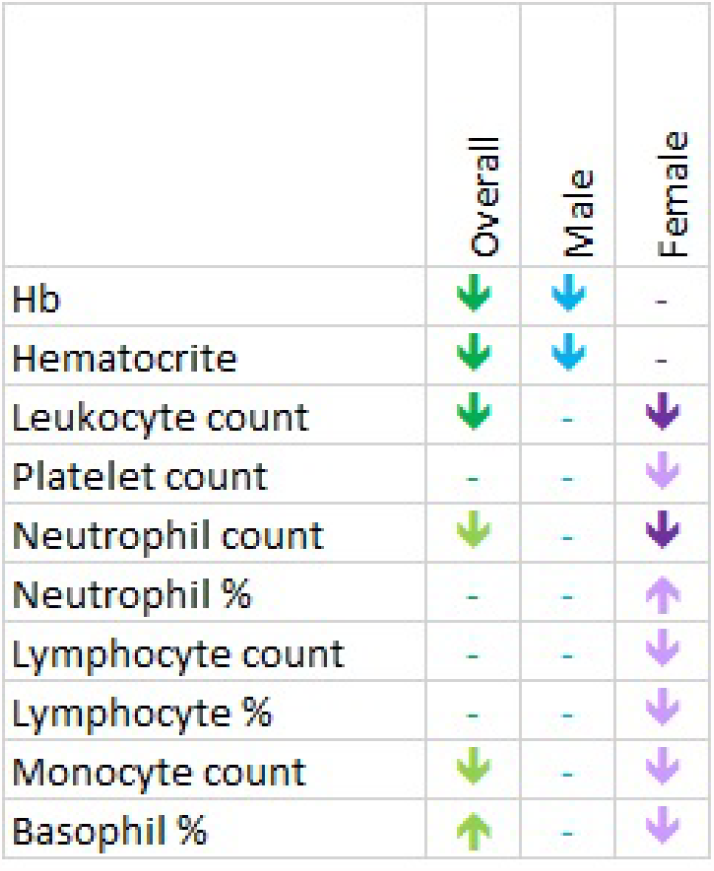
Summary of haematological features in SARS-CoV-2 positive donations as compared to negative ones. Intense-coloured arrows represent significant differences. Light-coloured arrows represent statistical trends.

## Discussion

A similar seroprevalence to that reported in general population was found among blood donors without COVID-19 antecedents. The 6.73% seroprevalence is a little lower than that reported in the seroprevalence study ENE-COVID-19 promoted by the *Instituto Nacional the Epidemiología* [11], where it is reported that a 7.2% of the participants in our region would have anti SARS-CoV2 IgG antibodies by the end of June 2020. Almost one quarter people in our region (25.51% according to National Institute of Statistics, INE 2020 data) is older than 65 and elderly are necessarily underrepresented in this study. That happens as well with population aged under 18 (<4.5% in our region). One limitation of this study is this of restricted age, but it would make just a little difference of 0.5% in seroprevalence estimation. Immunization rate would be a bit higher regarding the fact that some individuals who do not exhibit antibodies but still mount an efficient specific T-cell response [2]

Perhaps the main strength of our study is the large number of samples we randomized and the long period we tested (15 months). The first recorded COVID-19 case in Spain was reported on 31/01/2020, but three positive donations were found as testing samples collected along 2019. That could be meaningful in two opposite ways: one possible explanation would be cross reactivity to seasonal cold coronaviruses and the other would be that SARS-CoV-2 might have been circulating at least since 2019 summer.

It is now known that up to 28% people would have protection against SARS-CoV-2 due to cross reaction to other coronaviruses [2]. It has been checked that long-term cross-reactive both T-cells and antibodies can be correlates of protection against COVID-19 [12]. Asymptomatic or mild cases couldn’t be diagnosed or reported before the first tests were available. There are other reports about SARS-CoV-2 circulating in Europe along 2019 [13]. Our three 2019 positive donors would be even earlier cases, but the simplest explanation would be that of cross-reactions. Some of the positives along the period of study might be as well cross-reactive or both reactive due to SARS-CoV-2 infection together with previous coronavirus immunity.

Conversely to clinical forms of COVID-19 [14,15] neither age nor sex have an influence on the probability to develop an asymptomatic COVID-19 infection in our cohort. Several smaller seroprevalence reports from European countries support this feature [16-20]. Age and sex would not therefore have a role in SARSCoV-2 infection susceptibility, but only in their progression to severe forms.

Blood group has been reported elsewhere [7,21-23] to confere susceptibility or determine severity of SARS-CoV2 infection. Most published studies do not separate them. It should be noticed that there are no large studies characterizing asymptomatic cases in literature, perhaps this is one of the reports comprising one of the largest asymptomatic cohorts to date. Relationship of ABO system with susceptibility is not supported by our data. We excluded convalescent plasma donors, opposite to other published studies, that focused into these asymptomatic positive donors [22]. Perhaps blood group might play a role only in symptomatic cases and would be therefore related to severity but not to asymptomatic SARS-CoV-2 infection. Another surface antigens such as HLA or KIR might be as well involved and should be studied.

Several blood count anomalies have been reported in SARS-CoV2 infection, notably lymphopenia and neutrophilia are related to the severest forms [24]. Our data conversely reveal that asymptomatic positive women had a significant lower count of WBC and neutrophils as compared to negative ones. Similar low counts of WBC and neutrophils have been reported in young (<19) cohorts, comprising mainly mild and asymptomatic cases [25]. Perhaps this feature might be therefore a correlate of mildness. Elevations of monocytes have been elsewhere reported in short series of mild cases [26]. Our observation will be in the same line of those findings but a gender bias cannot be discarded so far seropositive women would go in the opposite direction.

Several reports about haemolytic anaemia secondary to SARSCoV-2 have been published [27-30]. We cannot discard a mild silent haemolytic anaemia in asymptomatic cases but just small asymptomatic cohorts are published to date.

We can conclude that seroprevalence estimations through blood donation analysis mirror population-based surveys. Sex and age would not affect COVID-19 susceptibility but its severity. Gender differences are present even in asymptomatic individuals: positive females have lower lymphocyte counts whereas positive males, present anaemia more frequently than negative ones. Further studies in large cohorts are needed to confirm these gender differences and to characterize asymptomatic COVID-19 cases as they can help better understand immune response to COVID-19, its pathogenesis and prognosis.

## Data Availability

Data collected for the study, including fully anonymized participant data, are available to others. Data available include fully anonymized participant data and data dictionary. Related documents are available from the date of publications henceforth: study protocol, statistical analysis, and approval of Ethical Board. These documents are available from the date of publications henceforth at email address. Data would be shared after approval of proposals by the Valladolid Este Ethical Committee.

## Acknowledgements

LB and MCM conceived and designed the study, NH, NO; MP and AP acquired data, IP, AJ and MIGF performed analysis and interpretation of laboratory data, MCM and LB drafted the article and revised it critically for important intellectual content, all authors provided final approval of the version to be submitted.

The authors thank to all blood donors for making this work possible by allowing research use of their samples by the Biobanco del Centro de Hemoterapia y Hemodonación de Castilla y León. They also thank the staff in charge of blood donation, and lab technicians for their efforts and Roche for its support.

## Abbreviations

(IgG): Immunoglobulin G
(IgA): immunoglobulin A
(IgM): immunoglobulin M
(PCR): polymerase chain reaction
(SD): standard deviation
(IQR): interquartile range
(WBC): White blood cells
(Hb): haemoglobin
(HCT): haematocrit

## Notes

### Competing Interest Statement

This work has been carried out provided free equipment and test reagents from Roche Diagnostics International Ltd. M Carmen Martin spoke about this issue in a sponsored symposium of Roche Diagnostics International Ltd.

### Funding Statement

This work has been carried out provided free equipment and test reagents from Roche Diagnostics International Ltd.

### Author Declarations

This study was conducted according with national regulations, institutional policies and in the tenets of the Helsinki Declaration. This study was approved, with the Valladolid Health Area Drug Research Ethics Committee acting as the main committee, in a meeting held on June 11th, 2020 with the reference number BIO-2020-93

### Summary of Updates

Baseline characteristics and data fron the original population added

